# Physician and Endoscopy Nurse Perspectives on Tap Water for Gastrointestinal Endoscopy: A Cross-Sectional Survey on Support, Perceived Barriers, and Intent to Advocate

**DOI:** 10.1101/2024.11.23.24317703

**Authors:** Anthony James Goodings, Allison Dana Chhor, Hannah Anderson, Mila Pastrak, Sten Kajitani, Matthew Schultzel, Aoife O’Sullivan, Ann-Marie Eustaceryan

## Abstract

**Background:** The standard water used for endoscopic irrigation is sterile water. Minimal evidence exists regarding sterile water use where there is access to clean water. The WHO has declared the climate crisis as the greatest global health crisis today; we must re-examine our practices and adapt them to promote environmental stewardship while maintaining safety.

**Method and Objective:** We surveyed physicians and endoscopy nurses to determine their attitudes toward tap water use for irrigation in gastrointestinal endoscopic procedures.

**Results:** There were 88 complete responses collected from June to November 2024. The majority of respondents and endoscopy-performing consultants expressed comfort with tap water use (59% and 84%, respectively), perceived viability (62% and 68%, respectively), and an interest to implement (73% and 94%, respectively); however, discussions on the topic remained infrequent (77% and 81%, respectively). 82% of overall respondents and 93% of consultants were aware of potential cost-savings, with 69% and 87% more willing to consider tap water based on this. Respondents (60%) and consultants (73%) agree there is a lack of guidelines regarding tap water use and feel that policy barriers will hinder change (59% and 73% respectively). Overall, 59% of respondents and 73% of consultants are likely to advocate for change.

**Conclusion:** The majority of respondents support tap water as a viable, cost-effective alternative with environmental benefits. A strong intention to advocate for change highlights the presence of potential leaders in this space. By promoting and supporting these leaders through education and institutional change, a more sustainable future for endoscopy exists.

**What is already known on this topic:**

- There is an urgent need to address the immense impact of medicine on the environment.
- Gastrointestinal endoscopy involves the use of large volumes of sterile water to irrigate a non-sterile space in the body.

**What this study adds:**

- Allows us to understand the views of physicians on the potential use of tap water in GI endoscopy, as well as understand perceived feasibility and barriers.

**How this study might affect research, practice, or policy:**

- With an understanding of physician support and perceived barriers, specific actions to address these can be taken by regulators.
- By increasing awareness around the topic, experts can deliberate on the idea and choose to advocate for change, providing leadership in an intimidating discipline.

## Introduction

Planetary health is the notion that the health of humans is dependent on the health of the natural environment. As physicians and leaders in our communities, we have a responsibility to advocate for the health of our patients, but also the health of future generations. This involves critically assessing our current practices and considering ways we can adapt to minimize our environmental impact, while preserving the high level of safety expected during medical interventions. The medical industry is a disproportionately large contributor to both greenhouse emissions and waste products^1,2^, particularly single-use plastics^3^. Health care emissions account for 8-10% of national emissions in the United States of America, 7% in Australia, and 5% in Canada^1^. Globally, healthcare represents up to 5% of total environmental impact^2^. Change is of paramount importance to minimize the negative impact climate change on human health^4^.

Gastroenterology is inherently a resource-intensive specialty, with studies identifying it as the third-highest contributor to waste in healthcare, largely due to endoscopy^5,6^. These procedures carry a high carbon footprint due to factors including the decontamination process, reliance on single-use products, medical waste production, and high case volume. A commonly overlooked aspect is the use of sterile water. A single endoscopic procedure utilizes up to 55 liters of water between decontamination of the endoscope and the use of irrigation water^7^. Much of this water is sterile, but we must question the rationale behind using sterile water. The gastrointestinal (GI) tract that is accessed during esophagogastroduodenoscopies (EGD) and colonoscopies is not sterile. We consume non-sterile water within these structures every day in the form of drinking water. This leads us and many of our colleagues^8,9^ to question the practice of sterile water use for non-sterile GI procedures (EGD and colonoscopies).

There has been recent work examining the general perceptions of endoscopists regarding sustainability in general. A 2024 survey of healthcare providers involved in endoscopy found that nearly 85% of respondents believe the British Society of Gastroenterology (BSG) should prioritize sustainability in GI endoscopy^10^.

Despite numerous professional societies recommending the use of sterile water in GI endoscopic procedures^11^, there is little to no evidence to demonstrate enhanced safety or improved patient outcomes. A small number of comparative studies over the past several decades have demonstrated that there is no difference in outcome between sterile versus tap water^12–14^. Of note, these studies were conducted in regions with access to potable tap water. Additionally, it is worth acknowledging that tap water is already used within the lower GI tract in the form of tap water enemas, with cohort studies reporting on the safety and efficacy of this practice^15,16^. In terms of colonoscopy, one scoping review found no adverse events associated with tap water for intraprocedural irrigation^14^.

Some articles comment on considering the use of tap water for GI endoscopy due to the lack of evidence of adverse clinical outcomes and environmental benefit^5,8^. However, little is known about the opinions of key clinical stakeholders on this issue. Relevant positions of the BSG indicate that tap water may be used for manual flushes through biopsy valves and that further research is needed into sustainable alternatives to sterile water in endoscopy suites^17^.

While there is no established difference in outcome, there are very important differences in environmental impact. The process of sterilizing, packaging, transporting, and disposing of the water and its packaging uses a significant amount energy and produces a large volume of waste. Although to our knowledge, there is no formal life cycle analysis of sterile water use in endoscopy, one study estimates the carbon footprint of endoscopy in the United States to be 85,768 metric tons of CO2 emissions annually based on energy usage and plastic waste alone, equivalent to burning 94 million pounds of coal^18^. A low-barrier and potentially high-yield way to mitigate this is replacing sterile water with tap water in endoscopy.

Municipal systems are already in place in developed countries to deliver potable water to every household and hospital. The environmental impact of switching from sterile water stored in single-use plastic containers to tap water for non-sterile endoscopic procedures is significant, warranting a reconsideration of current practices.

For changes to be implemented, a thorough understanding of the attitudes of endoscopists towards the issue is necessary to foster leadership, build advocacy, and drive behavioral change. Therefore, pursuing quality improvement studies^19^ and understanding the opinions of key stakeholders is essential in moving towards a sustainable future in endoscopy.

### Objectives

1. Understand the attitudes of endoscopists towards the use of tap-water for endoscopic irrigation.
2. Understand the role of environmental and cost-based considerations on willingness to change practice.
3. Understand the barriers to implementing sustainable changes in endoscopic medicine.

## Methods

### Inclusion Criteria and Study Population

We performed an observational study of physician and endoscopy nurses. Data pertaining to participant specialty and involvement in GI endoscopic procedures were obtained for the purpose of forming distinct subgroups. Data collection took place from June to November 2024.

### Data Collection

Eligible individuals were contacted directly, via colleagues, and via professional organizations (e.g. Irish Society of Gastroenterology) with a link to complete the online survey. Distribution on publicly accessible social media websites was not conducted to preserve data integrity. The survey was hosted on the *Qualtrics* professional survey platform.

### Key Outcome Measures

The primary outcome is to determine the attitudes of endoscopist towards tap water use in endoscopy across four principal domains: General Attitudes and Perceptions, Environmental Considerations, Cost Considerations, and Policy Impact. These are presented in *Table 1*. Secondary outcomes included understanding the general opinions in these domains across all physicians, trainees, and endoscopic nurses.

**Table 1:** Survey Questions.

| General Attitudes and Perceptions |
| --- |
| I would be comfortable using tap water for endoscopic irrigation. |
| I have safety concerns in using tap water for endoscopic procedures. |
| I believe that tap water would be as effective as sterile water during endoscopic procedures. |
| Tap water is a viable alternative to sterile water for endoscopic irrigation in centers with access to clean water. |
| I believe that the safety of using tap water compared to sterile water for endoscopic irrigation is equivalent. |
| How often do you encounter discussions or considerations regarding the choice between tap water and sterile water for endoscopic irrigation in your clinical practice? |
| I trust the recommendations provided by professional organizations regarding the use of tap water for endoscopic procedures. |
| How comfortable are you discussing the use of tap water for endoscopic procedures with patients or their caregivers? |
| I am interested in implementing tap water irrigation in my practice. |
| Environmental Considerations |
| How important is it to minimize the environmental footprint of medical procedures, such as endoscopy? |
| I am aware of the environmental impact associated with the use of sterile water in plastic containers for endoscopy. |
| To what extent would environmental considerations influence your decision to use tap water instead of sterile water in endoscopic procedures? |
| How important do you believe it is that healthcare facilities should prioritize environmentally friendly practices, such as using tap water instead of sterile water, in their daily operations? |
| I have a personal responsibility to contribute towards sustainable healthcare by engaging in sustainable practices. |
| It is reasonable to consider changing your practice in the absence of randomized control trial evidence, when there is no reason to suspect patient harm, for the benefit of the environment. |
| How likely are you to advocate for environmentally friendly practices within your healthcare facility, such as using tap water for endoscopic irrigation? |
| **Cost Consideration** |
| I am aware of the cost differences between using tap water and sterile water for endoscopic procedures. |
| How significant is the cost in your decision-making process regarding which products are used for endoscopic procedures? |
| My workplace takes into account cost effectiveness when choosing which products are used for endoscopic procedures. |
| Using tap water instead of sterile water for endoscopic procedures could lead to significant cost savings. |
| The potential cost savings associated with using tap water would impact my willingness to consider it as an alternative to sterile water. |
| I would use the narrative of cost saving to advocate for the adoption of tap water irrigation in my center. |
| **Policy Impact** |
| I have confidence that guidelines for endoscopic procedures are based on strong scientific evidence. |
| I am aware of which regulatory body created the guidelines for endoscopic procedures used in facility. |
| I am aware of evidence behind my healthcare facilities' current guidelines in regards to what is used during endoscopic procedures. |
| There are a lack of clear guidelines regarding tap water use that impacts my willingness to consider it as an alternative to sterile water. |
| To what extent do you perceive regulatory/policy barriers would hinder the widespread adoption of tap water for endoscopic irrigation? |
| How likely are you to advocate for policy changes to encourage the use of the tap water for endoscopic irrigation? |

### Statistical Analysis

Descriptive statistics were performed using SPSS V.28.

## Results

### Study Cohort

There was a total of 88 complete responses, these were included in the analysis. In the case of missing response for a single question, that respondent was omitted from the tally for that specific question.

The principal subgroup was *endoscopy-performing consultant/attending physicians* of which there were 31 responses (27 gastroenterologists, 3 general surgeons, 1 colorectal surgeon). The remainder of respondents consisted of endoscopy nurses (36), specialist registrars / residents in a medical specialty (11), other non-consultant hospital doctors (5), other consultant/attendings (3), specialist registrars / residents in a surgical specialty (2).

Respondents were primarily from Europe/UK (78), with some from North America (7), and some from other regions (3).

Respondents’ gender identity was 41% male (n = 36) and 59% female (n = 52).

### General Attitudes and Perceptions

There was a generally positive response to the questions regarding general attitudes towards tap-water use. Consultants were generally in stronger agreement than overall respondents. These can be visualized in *Figure 1* and are summarized in the following subsections.

**Figure 1.**
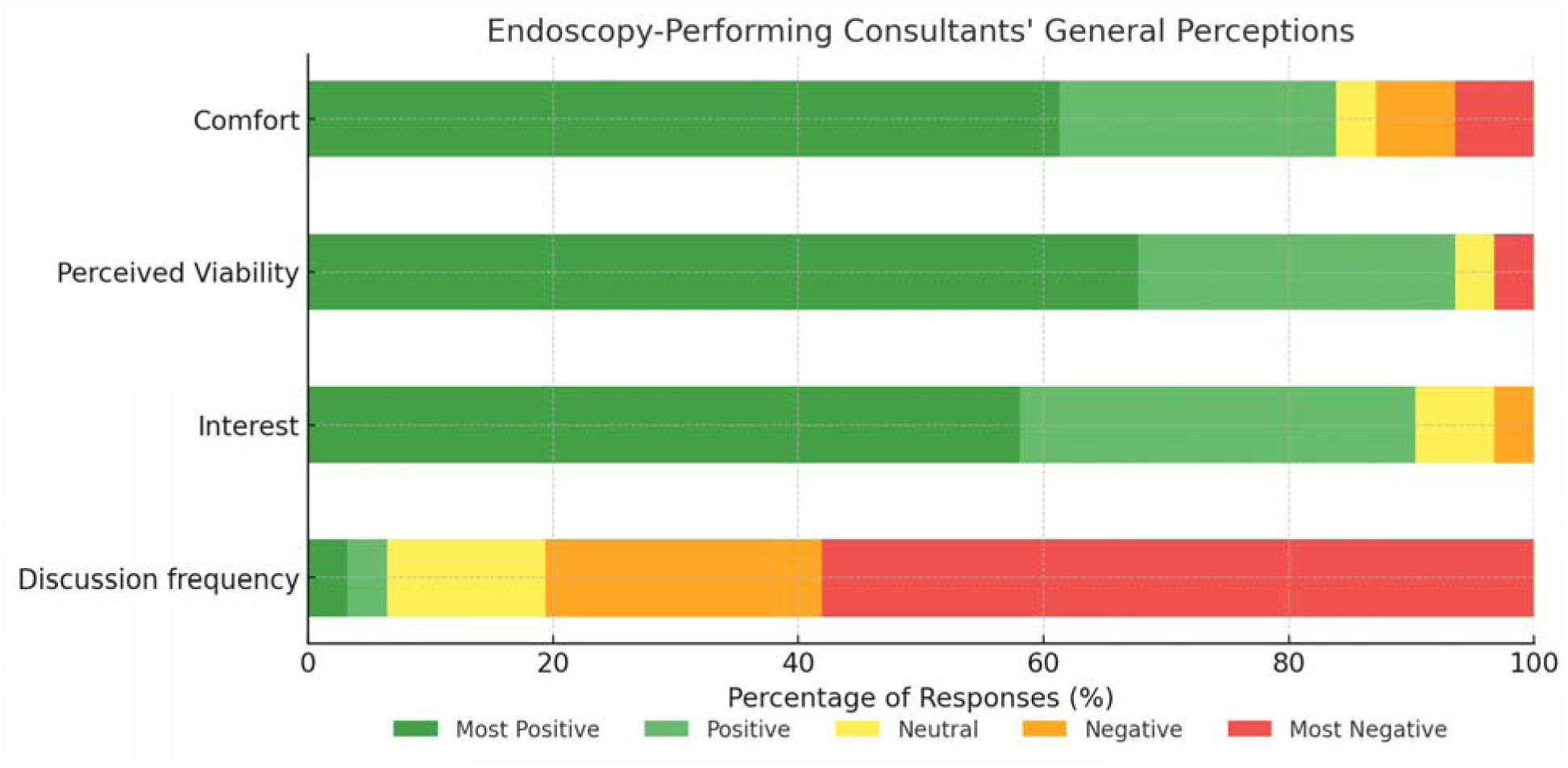
Responses of endoscopy-performing consultants regarding tap-water use

#### Comfort with Tap Water User

Respondents indicated agreement (35% strongly, 24% somewhat) and among endoscopy-performing consultants the agreement was stronger (61% strongly, 23% somewhat).

#### Perceived Viability

Overall, respondents (46% strongly, 27% somewhat) and endoscopy-performing consultants (68% strongly, 26% somewhat) agreed that tap water is a viable alternative to sterile water for irrigation.

#### Interest in Implementation

There was strong interest in implementing tap water, 37% of all respondents and 58% of endoscopy-performing consultants were strongly interested. Additionally, 33% of both groups were somewhat interested.

#### Frequency of Discussion

Discussion of tap-water endoscopy was rare. 58% of all respondents and consultants indicated that the topic was never discussed and 19% and 23% indicated that it was rarely discussed respectively in their workplace.

### Environmental Considerations

Responses reflected strong interest, and a high degree of importance attributed to environmentally friendly practices. These results can be visualized in *Figure 2*.

**Figure 2.**
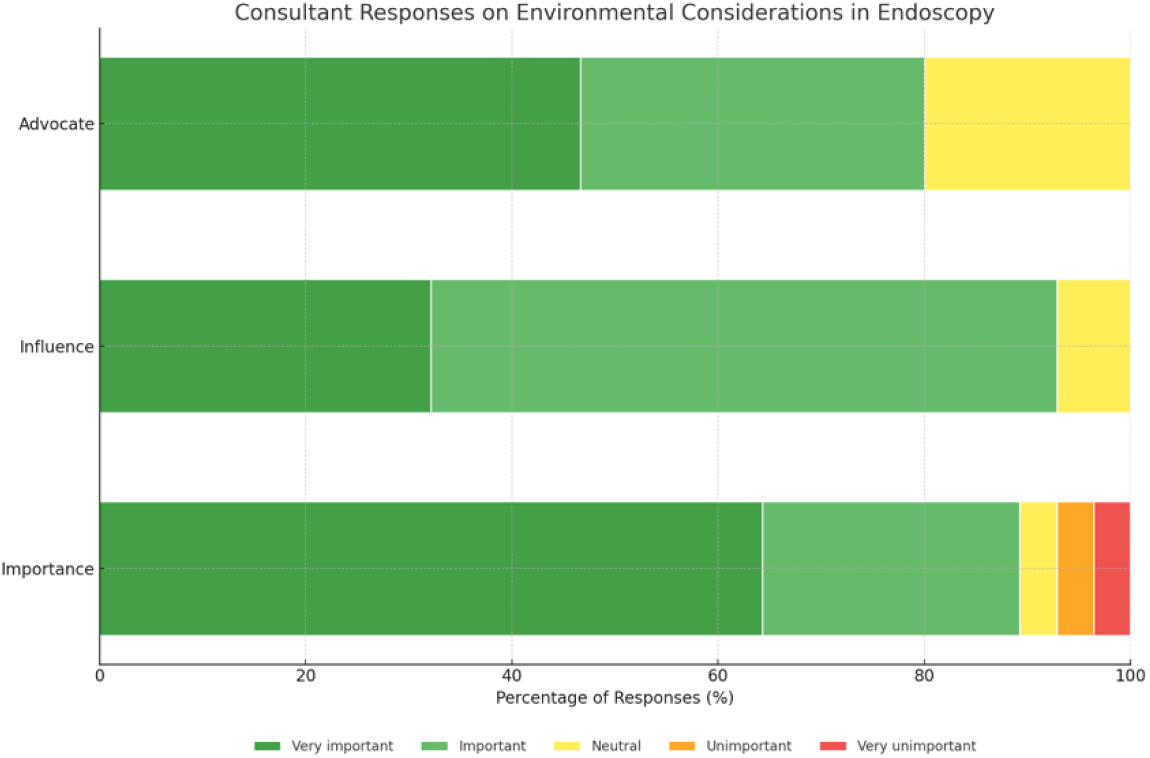
Environmental considerations in endoscopy

#### Importance of Minimizing Environmental Footprint

Most respondents found it very important (53%) or important (26%) to minimize the environmental impact of procedures. Among endoscopy-performing consultants it was 64% and 25% respectively.

#### Influence of Environmental Factors on Decision-Making

Among all respondents, 27% and 42% reported that environmental factors play a large or very large role in their decision-making regarding the type of water used, respectively. Among consultants, the role of the environment was more prominent, with 32% reporting it plays a very large role and 61% saying it plays a large role.

#### Likelihood to Advocate for Environmentally Friendly Practices

For all respondents, 31% were very likely and an additional 34% were likely to advocate for environmentally friendly practices in their workplace. Among endoscopy-performing consultants, 47% were very likely to do so and 33% were somewhat likely to.

### Cost Considerations

Responses reflected general agreement regarding the importance of cost. The results are visualized in *Figure 3*.

**Figure 3.**
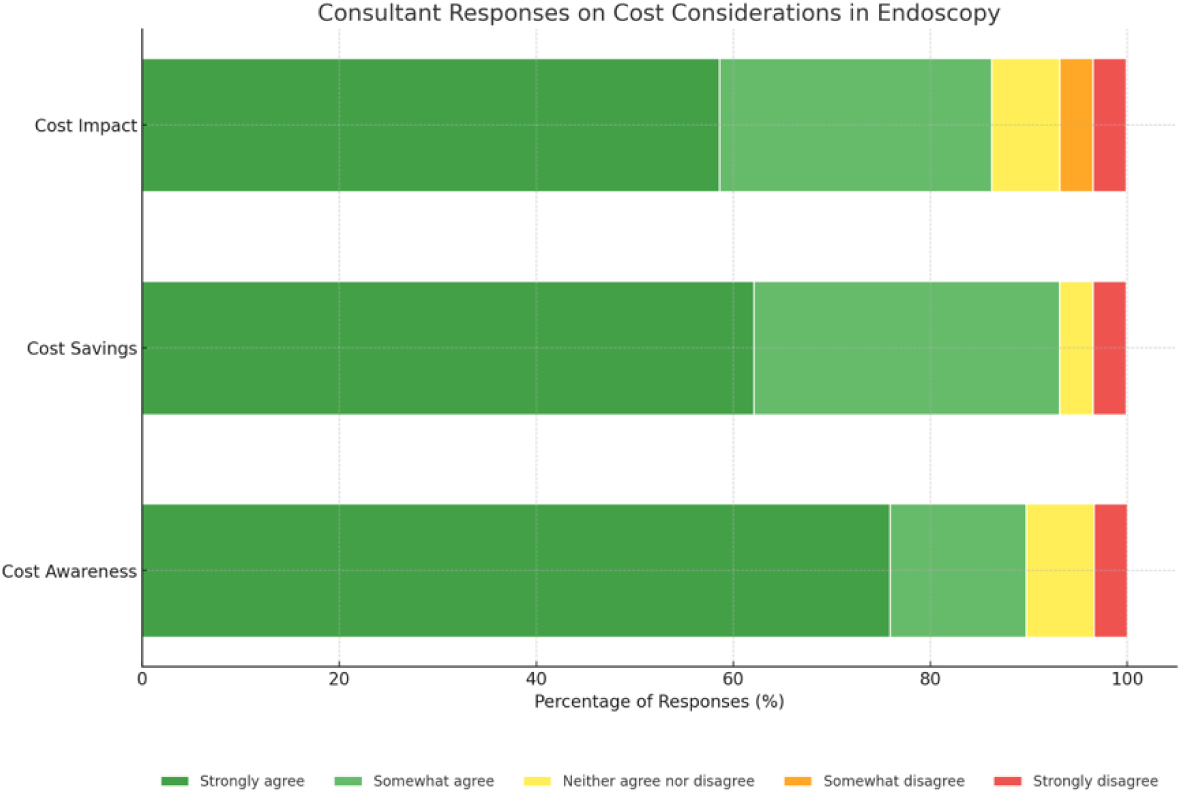
Cost considerations relating to tap water use in endoscopy

**Figure 4.**
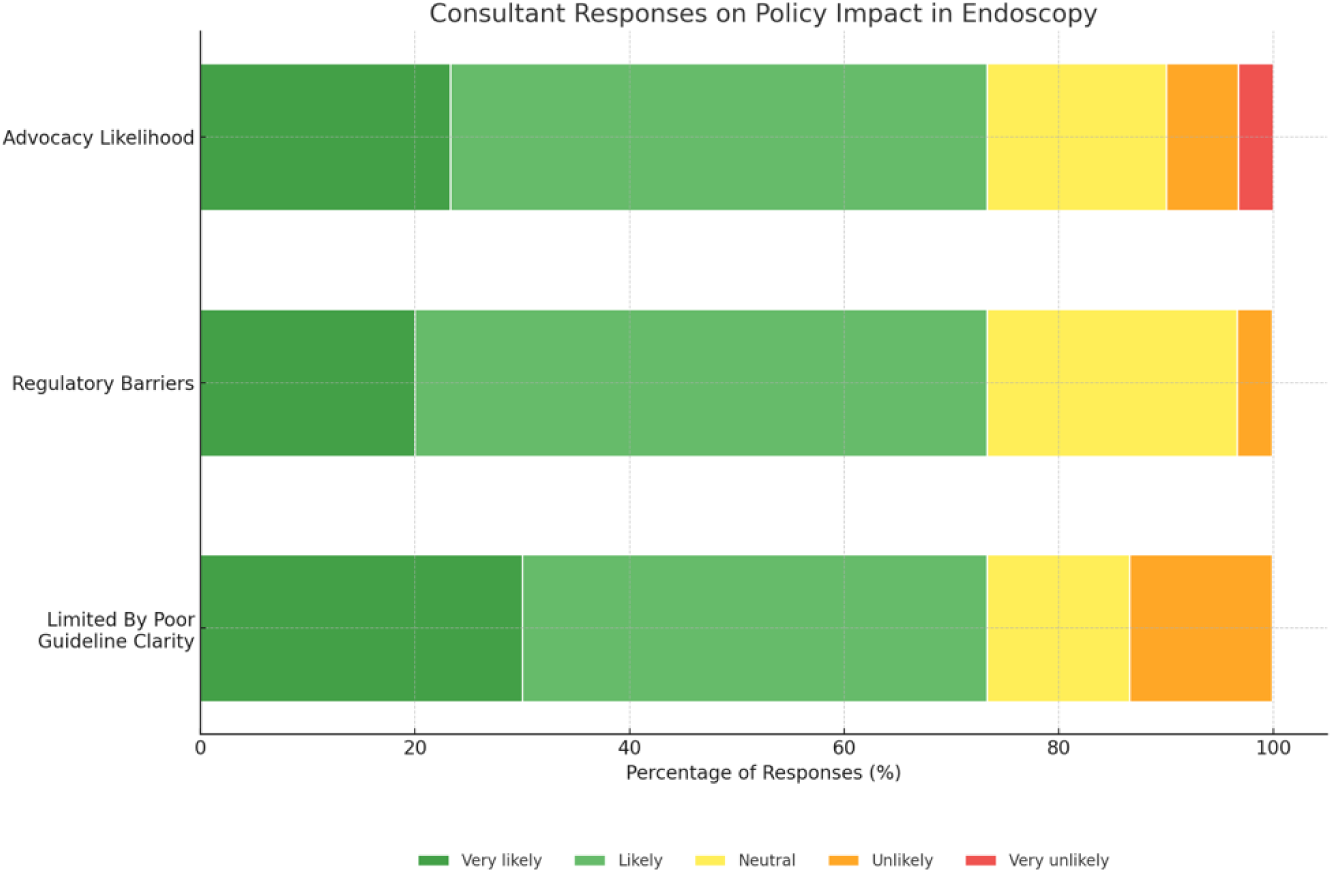
Likelihood to advocate and key limitations to adoption

#### Awareness of Cost Differences

Of all respondents 47% indicated strong agreement regarding knowledge of the cost difference between methods, 20% somewhat agreed. Among endoscopy-performing consultants, 76% and 14% indicated the same.

#### Potential for Cost Savings

51% of respondents strongly agreed and 31% somewhat agreed that using tap water would lead to cost savings. Among endoscopy-performing consultants 62% and 31% felt the same respectively.

#### Impact of Cost Savings on Willingness to Consider the Use of Tap Water

43% of respondents agreed strongly while 26% agreed somewhat that the cost-saving impacts their willingness to use tap water. Among the endoscopy-performing consultants 59% agreed strongly while 28% agreed somewhat.

### Policy Impact

#### Impact of Lack of Clear Guidelines Regarding Tap Water Use

Overall, 30% strongly agreed and an additional 30% somewhat agreed that a lack of clear guidelines impacts their consideration of tap water use. Among endoscopy-performing consultants 30% strongly agreed and 43% somewhat agreed with the same.

#### Perceived Policy Barriers

Of all respondents 18% and 41% perceive that policy barriers would, to a very large and large extent respectively, hinder the widespread adoption of tap water use in endoscopic irrigation. 20% and 53% of endoscopy-performing consultants felt the same.

#### Likelihood to Advocate for Tap Water Policy Change

59% of all respondents indicated they would be likely or very likely to advocate for policy change around the use of tap water for endoscopic irrigation. Among endoscopy-performing consultants 73% indicated the same.

### Changing Practice

Respondents were asked whether they agreed or disagreed with the following statement: “It is reasonable to consider changing your practice in the absence of randomized control trial evidence, when there is no reason to suspect patient harm, for the benefit of the environment.” Among endoscopy-performing consultants, 39% strongly agreed, 7% somewhat agreed, 25% somewhat disagreed, and 29% strongly disagreed.

## Discussion

In this cross-sectional study, we surveyed physicians and endoscopy nurses to assess their perception and consideration of the environmental impact, cost, and policies regarding tap water use in endoscopic procedures. Over half of our respondents were endoscopic-performing consultants across various hospitals, whose expertise in the field contributes to the impact of their views. We found that the majority of these consultants felt comfortable using tap water irrigation, believe it to be a viable alternative, and were interested in implementing this into their practice. Most respondents believe it is important to minimize the environmental impact of medical procedures such as endoscopy, and that this impact plays a large role in their choice of the type of water used. This support for sustainable practice is in line with the growing awareness and advocacy for sustainability in healthcare worldwide^20^. Our findings, showing strong physician support for tap water use in endoscopy, help to make the case for its safety using first principles. Overall, among endoscopy-performing consultants surveyed in this study, the concept of changing practice when there is no reason to suspect patient harm without evidence from randomized control trials is very contentious, with a near-even split in opinion between experts. The intent to advocate to other healthcare professionals and patients can progress the advancement of sustainable medicine. As physicians are a trusted and credible voice in our society, further advocacy in this area can help to promote acceptance and uptake of this practice amongst endoscopists. An initial adoption coupled with robust reporting of outcomes may allow for professionals to feel more comfortable in adopting tap water as a sustainable alternative.

In order to establish progress in this discipline, strong physician and institutional leaders are essential. There is a strong case for every institution to have a director of planetary health^21,21^. Senior leadership can significantly impact junior trainees and foster a culture of openness and environmental prioritization when appropriate, this is evident through faculty supported, student led initiatives such as the Planetary Health Report Card^22^.

In terms of cost, most respondents were aware of the potential cost differences between tap and sterile water, agreeing that changing to tap water would lead to cost savings, and that this would subsequently lead to willingness to switch methods. This aligns with the Sustainability in Quality Improvement (SusQI) framework, which outlines that measuring the impact and return on investment of an action helps drive change by communicating its benefit to others^23^. Further studies that quantify the financial cost of sterile water used in endoscopy suites could help to promote the adoption of tap water where appropriate, encouraging administrators and policy makers to consider tap water as a cost-effective, safe alternative.

Approximately half of respondents agreed that a lack of clear guidelines impacts their consideration of tap water use, and that policy barriers would hinder the widespread adoption of tap water use in endoscopic procedures. The development of evidence-based guidelines from reputable international bodies could foster increased confidence amongst clinicians, encouraging broader adoption of this practice.

The majority of respondents indicated they were likely to advocate for environmentally friendly practices in their workplace, as well as policy change around the use of tap water for endoscopic irrigation. In a recent cross-sectional survey of UK doctors, approximately 89% supported policy and guideline development on sustainable endoscopy practice^10^. Our study reaffirms these sentiments, demonstrating strong support and willingness to drive change in this area. A comprehensive life cycle analysis of sterile water use in endoscopy would provide compelling data to aid advocacy efforts.

Limitations of the study include the regional demographics, with 85% of respondents being from Ireland or the UK, potentially limiting generalizability and reflecting the opinions of physicians primarily working in a public healthcare system. Additionally, there exists a self-selection bias, as physicians who are more interested in sustainability may have been more likely to opt in to completing the survey. This may lead to an overrepresentation of physicians who are more likely to support sustainable healthcare practices. Lastly, we were not able to explore the reasons why some physicians were not interested in using tap water at an individual level. Further studies could include focus groups to address these aspects, offering valuable insight into physicians’ concerns to inform the development of practical, sustainable solutions.

Our work suggests not only the need, but the desire for clinical studies in this area. Cohort or randomized controlled trials can lead to policy changes that allow endoscopic providers to change their practice. Furthermore, this study can promote future investigations into how medical practices impact the environment, and strategies to limit this impact. This is with the ultimate goal of developing strong leaders in medicine who will advocate for institutional change in our evolving world.

## Summary

The findings of this study revealed a significant level of interest among physicians in adopting tap water for endoscopic irrigation, underscoring its perceived viability as an alternative. The is also a strong interest in advocacy, particularly in senior physicians, this provides an understanding of the presence of potential leaders and the need to support them. However, the absence of clear evidence-based guidelines and the perceived presence of policy barriers were identified as major obstacles preventing its use. Addressing these challenges could pave the way for more sustainable endoscopic practices, offering both environmental and clinical benefits.

## Data Availability

All data produced in the present study are available upon reasonable request to the authors.

## Ethics Statement

Project approved by the Social Research Ethics Committee, University College Cork. Reference number: SOM/SREC/2024/2103/1

## Funding

This project received no specific funding.

## Competing Interest Statement

The authors have no competing interests to declare.

## Notes

### Competing Interest Statement

The authors have declared no competing interest.

### Funding Statement

This study did not receive any funding.

### Author Declarations

The Social Research Ethics Committee of University College Cork gave ethical approval for this work.

### Summary of Updates

Elaborate on themes of Advocacy and Leadership.

